# UBR1 Promotes Sex-Dependent ACE2 Ubiquitination in Hypertension

**DOI:** 10.1101/2024.05.23.24307722

**Authors:** Mona Elgazzaz, Navya Lakkappa, Clara Berdasco, Uma Priya Mohan, Anna Nuzzo, Luke Restivo, Alexa Martinez, Amy Scarborough, Jessie J. Guidry, Srinivas Sriramula, Jiaxi Xu, Hisham Daoud, Michelle A. Mendiola Plá, Dawn E. Bowles, Andreas M. Beyer, Franck Mauvais-Jarvis, Xinping Yue, Catalin M. Filipeanu, Eric Lazartigues

## Abstract

**Background:** Angiotensin (Ang)-II impairs the function of the antihypertensive enzyme ACE2 by promoting its internalization, ubiquitination and degradation thus contributing to hypertension. However, few ACE2 ubiquitination partners have been identified and their role in hypertension remains unknown.

**Methods:** Proteomics and bioinformatic analysis were used to identify ACE2 ubiquitination partners in the brain, heart, and kidney from Ang-II-infused C57BL6/J mice from both sexes and validated the interaction between UBR1 and ACE2 in cells. Central and peripheral UBR1 knockdown was then performed in male mice to investigate its role in the maintenance of hypertension.

**Results:** Proteomics analysis from hypothalamus identified UBR1 as a potential E3 ligase promoting ACE2 ubiquitination. Enhanced UBR1 expression, associated with ACE2 reduction, was confirmed in various tissues from hypertensive male mice and human samples. Treatment of endothelial and smooth muscle cells with testosterone, but not 17β-estradiol, confirmed a sex-specific regulation of UBR1. In vivo silencing of UBR1 using chronic administration of small interference RNA resulted in the restoration of ACE2 levels in hypertensive males. A transient decrease in blood pressure following intracerebroventricular, but not systemic, infusion was also observed. Interestingly, UBR1 knockdown increased the brain activation of Nedd4-2, an E3 ligase promoting ACE2 ubiquitination and reduced expression of SGK1, the kinase inactivating Nedd4-2. Conclusions: These data demonstrate that UBR1 is a novel ubiquitin ligase targeting ACE2 in hypertension. UBR1 and Nedd4-2 E3 ligases appear to work synergistically to ubiquitinate ACE2. Targeting of these ubiquitin ligases may represent a novel strategy to restore ACE2 compensatory activity in hypertension.

## Introduction

Angiotensin Converting Enzyme 2 (ACE2) is an important compensatory enzyme of the renin-angiotensin system (RAS), converting the potent vasoconstrictor Angiotensin (Ang)-II to the vasodilatory peptide Ang-(1-7).^1^ We previously reported that ACE2 is the target of Ang-II-induced internalization and degradation via the Ang-II type 1 receptor (AT_1_R), a process involving ubiquitination.^2–4^ Ubiquitination is a three-step enzymatic process involving ubiquitin-activating enzymes (E1) binding to ubiquitin using a molecule of ATP, followed by ubiquitin-conjugating enzymes (E2), carrying the ubiquitin protein and conjugating with ubiquitin protein ligases (E3). E3 recognizes the protein to be labeled and catalyzes the transfer of ubiquitin to the target protein. The pathway continues until the target protein has a desired chain of poly-ubiquitins which is then subjected to lysosomal/proteasomal degradation.^5,6^ Ubiquitination plays an important role in the development of hypertension.^7^ Indeed, Nedd4-2, that we recently implicated in ACE2 ubiquitination,^3^ is a well-known E3 ligase controlling cell surface expression of kidney epithelial Na^+^ channels (ENaC) via ubiquitin-mediated endocytosis and lysosomal degradation.^7^ Mutations of ENaC have also been reported to interfere with ubiquitination by Nedd4-2, resulting in Liddle syndrome, a monogenic hypertensive condition.^8,9^ Expression of Nedd4-2 was enhanced in the hypothalamus of mice and endothelial cell cultures following Ang-II treatment and associated with a decrease in ACE2 while knockdown of Nedd4-2 restored ACE2 expression.^3^ Importantly, prevention of ACE2 ubiquitination resulted in the maintenance of a GABAergic inhibitory input to presympathetic neurons in the hypothalamus and a reduction of hypertension.

While our group was first to report that Ang-II treatment induces ACE2 internalization and ubiquitination through an AT_1_R-dependent mechanism,^2,4^ very few reports have focused on ACE2 ubiquitination partners.^1^ Murine double minute 2 (MDM2) E3 ligase was the first enzyme shown to promote ACE2 ubiquitination, possibly contributing to the development of pulmonary arterial hypertension.^10^ Another E3 ligase, S-Phase Kinase Associated Protein 2 (Skp2), was reported to be upregulated by cigarette smoke carcinogens, leading to ACE2 ubiquitination in lung epithelial cells.^11,12^ However, despite more than two decades of research establishing ACE2 as a critical player for cardiovascular regulation, and several recent studies related to SARS-CoV-2 infection,^1^ there is limited knowledge about the enzymes and mechanisms controlling ACE2 ubiquitination and the overall implications for cardiovascular regulation.

Incertitude also persists regarding sex differences in ACE2 expression and activity. Early work in rodents showed that ACE2 expression^13,14^ and activity^15^ are tissue-specific, higher in male kidneys and under the control of estrogens, while others reported no sex difference. Estrogens and testosterone are well known to affect the expression and activity of RAS components, contributing to sex differences in cardiovascular outcomes.^16,17^ Estrogens have also been reported to upregulate the protective Ang-(1-7)/ACE2/Mas1R/AT_2_R,^18^ notably by increasing ACE2 gene expression.^19^ In humans, studies have shown differential smooth muscle cells expression of ACE2 in the airways of males and females. Clinical studies also reported that higher levels of circulating soluble ACE2, previously shown by us to be a marker of disease,^20,21^ were associated with the male sex, cardiovascular disease, diabetes, and age; although increased ACE2 shedding does not imply sex differences at baseline.^22,23^ Hence, sex differences in ACE2 expression are still poorly understood and the effect of sex hormones on ACE2 ubiquitination in normotensive and hypertensive conditions has not been investigated.

In the current study, we identified UBR1, an E3 ligase part of the N-end rule proteolytic pathway of the ubiquitin system, as a sex-dependent regulator of ACE2 expression in hypertension. We further report that the knockdown of UBR1 prevented ACE2 degradation, which supports its role in ACE2 ubiquitination and could lay the foundation for new therapeutic strategies to prevent the loss of ACE2 compensatory activity in hypertension.

## Methods

The authors declare that all supporting data are available within the article and in the Data Supplement. A detailed Methods section is available in the Online Data Supplement.

Experiments were conducted in adult C57BL6/J mice (8–12 weeks old, 20-25 g; Jackson Laboratory, Bar Harbor, ME) from both sexes. Mice were housed in a temperature-(∼25 °C) and humidity-controlled facility under a 12-hour dark/light cycle, fed standard mouse chow (Envigo, iOS Teklab Extruded Rodent Diet 2019S, Huntingdon, UK) and water *ad libitum*. All procedures conformed to the National Institutes of Health Guide for the Care and Use of Laboratory Animals and were approved by the Louisiana State University Health Sciences Center (#3540), and the Southeast Veterans Healthcare System (#620) Institutional Animal Care and Use Committees. Cardiac samples were obtained from The Medical College of Wisconsin (IRB # PRO00010828) and Duke University (IRB # PRO00005621).

## Results

### UBR1 Ubiquitin ligase is predicted to interact with ACE2 in hypertension

To identify ACE2 ubiquitination partners in hypertension, a proteomic analysis was initially performed as reported previously.^24^ Following 4 weeks of Ang-II infusion, the hypothalamus was isolated and underwent proteomic analysis using Liquid chromatography-mass spectrometry (LC-MS) with a focus on ubiquitination-related proteins (Figure 1A). Based on abundance score, our data show an upregulation of the E3 ligase UBR1 in Ang-II-induced hypertension in wild type mice hypothalamus (Figure 1B and Table S1). To investigate whether ACE2 could be a potential target for UBR1, we used bioinformatic tools to evaluate the interaction between ACE2 and UBR1. Using the Biological General Repository for Interaction Datasets (BioGRID) to explore curated interactions between ACE2 and ubiquitin ligases,^25^ multiple ubiquitin ligases were shown to interact with ACE2 in humans, among which is UBR1 (Figure 1C).

**Figure 1:**
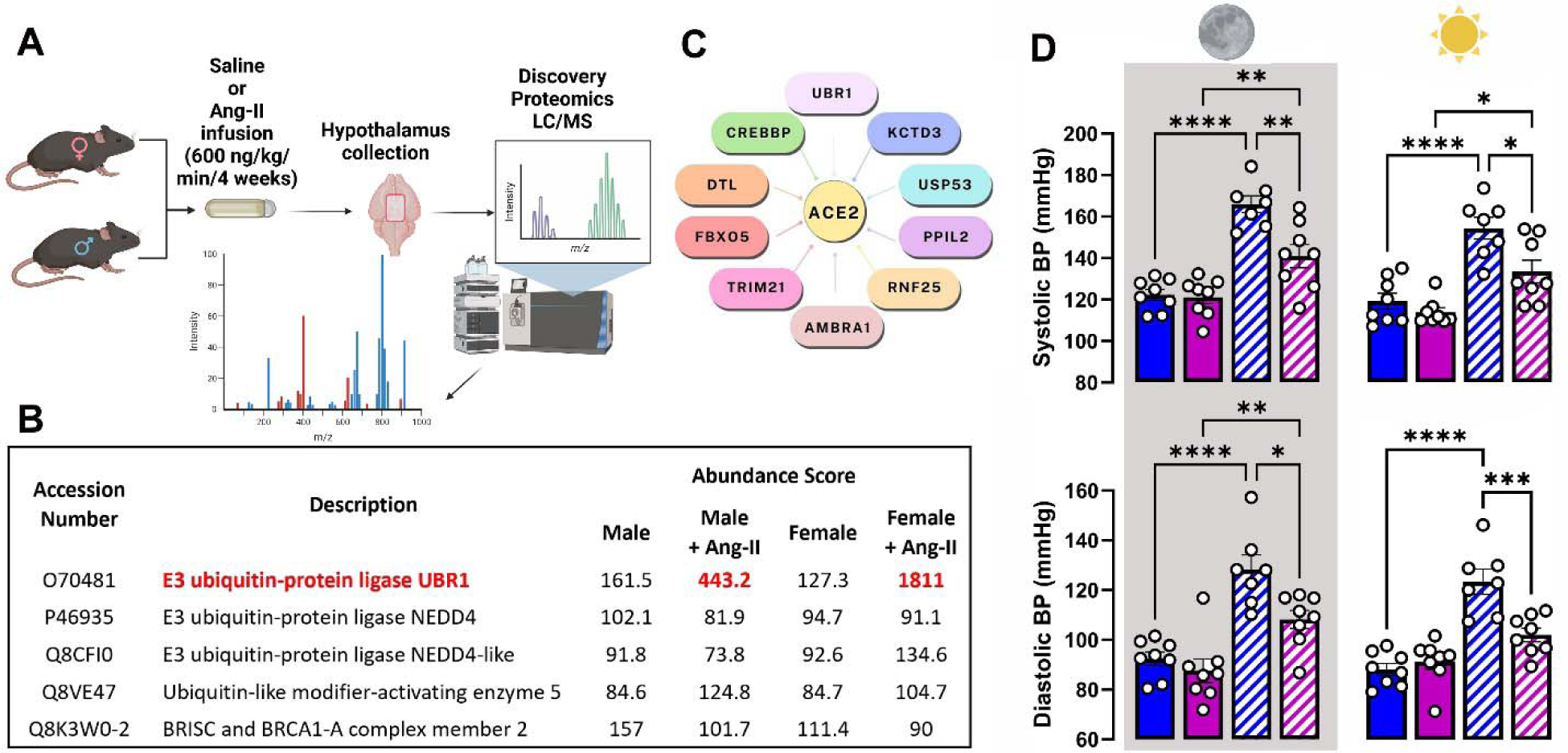
UBR1 is upregulated in hypertension and interacts with ACE2. (A) Hypothalami collected from sham- or Ang-II-treated C57Bl6/J male and female mice were subjected to proteomic analysis using Liquid chromatography–mass spectrometry (LC-MS). (B) Proteomics abundance scores of ubiquitin ligase proteins measured by LC-MS in hypertensive and normotensive mice. (C) Ubiquitin ligase system proteins interacting with ACE2 according to The Biological General Repository for Interaction Datasets (BioGRID; https://thebiogrid.org/ last accessed on February 10, 2024), showing ACE2 interacting proteins belonging to the ubiquitin proteasome system. (D) Systolic and diastolic BP were recorded in C57Bl6/J male (blue) and female (magenta) mice at baseline (solid bars) and four weeks after Ang-II (hatched bars) infusion (n=7-8/group) during the active (moon icon and shaded area) and resting phases (sun icon). Data are shown as mean ±SEM. Statistical significance: Two-way analysis of variance (ANOVA): *P<0.05, **P<0.01, ***P<0.001 and ****P<0.0001 *vs.* respective controls.

ACE2 ubiquitination was further investigated in hypertension induced by Ang-II infusion in both male and female mice. There was no significant blood pressure (BP) or heart rate difference between male and female mice before Ang-II treatment (Figure 1D and S1). Ang-II treatment resulted in a significant rise in BP that plateaued by the 4^th^ week of infusion. Hypertension was significantly higher in males than in females in both active and resting phases (Figure 1D).

### Sex hormones regulate the expression of UBR1 and ACE2

Protein expression for UBR1 and ACE2 was assessed in the mouse brain (Figure 2A), heart (Figure 2B) and kidneys (Figure 2C). We observed that male mice treated with Ang-II showed a 2 to 3-fold increase in UBR1 levels in those tissues. Although UBR1 expression trended to an increase in females, it only reached significance in males. ACE2 protein expression was significantly higher in males, notably in the brain (Figure 2E), while similar in the kidney for both sexes (Figure 2G). Although these higher levels were not significant in the mouse heart (Figure 2F), data from human left ventricles (Figure 2H) confirmed the elevated level in males. Ang-II treatment dramatically reduced ACE2 expression, notably in the male brain (Figure 2E) with limited (Figure 2G) or no impact (Figure 2E,F) in females. Together, these data suggest that Ang-II-induced upregulation of UBR1 in males could contribute to the downregulation of ACE2. To assess the clinical relevance of UBR1 in human hypertension, we evaluated left ventricle cardiac samples from rejected donor hearts (Males: 47.5 ±2.5, Females: 43.8 ±2.1 years old, n=6/group, Table S2) with or without hypertension. As observed in mice, ACE2 expression was higher in male normotensive donors (Figure 2H) and decreased with hypertension. UBR1 protein levels were strongly upregulated in hypertensive donors (Figure 2D), regardless of sex, but were associated with a decrease in ACE2 protein in males only (Figure 2H). Together, these data confirm that UBR1 upregulation and the concomitant ACE2 downregulation observed in our preclinical model is recapitulated in hypertensive subjects.

**Figure 2:**
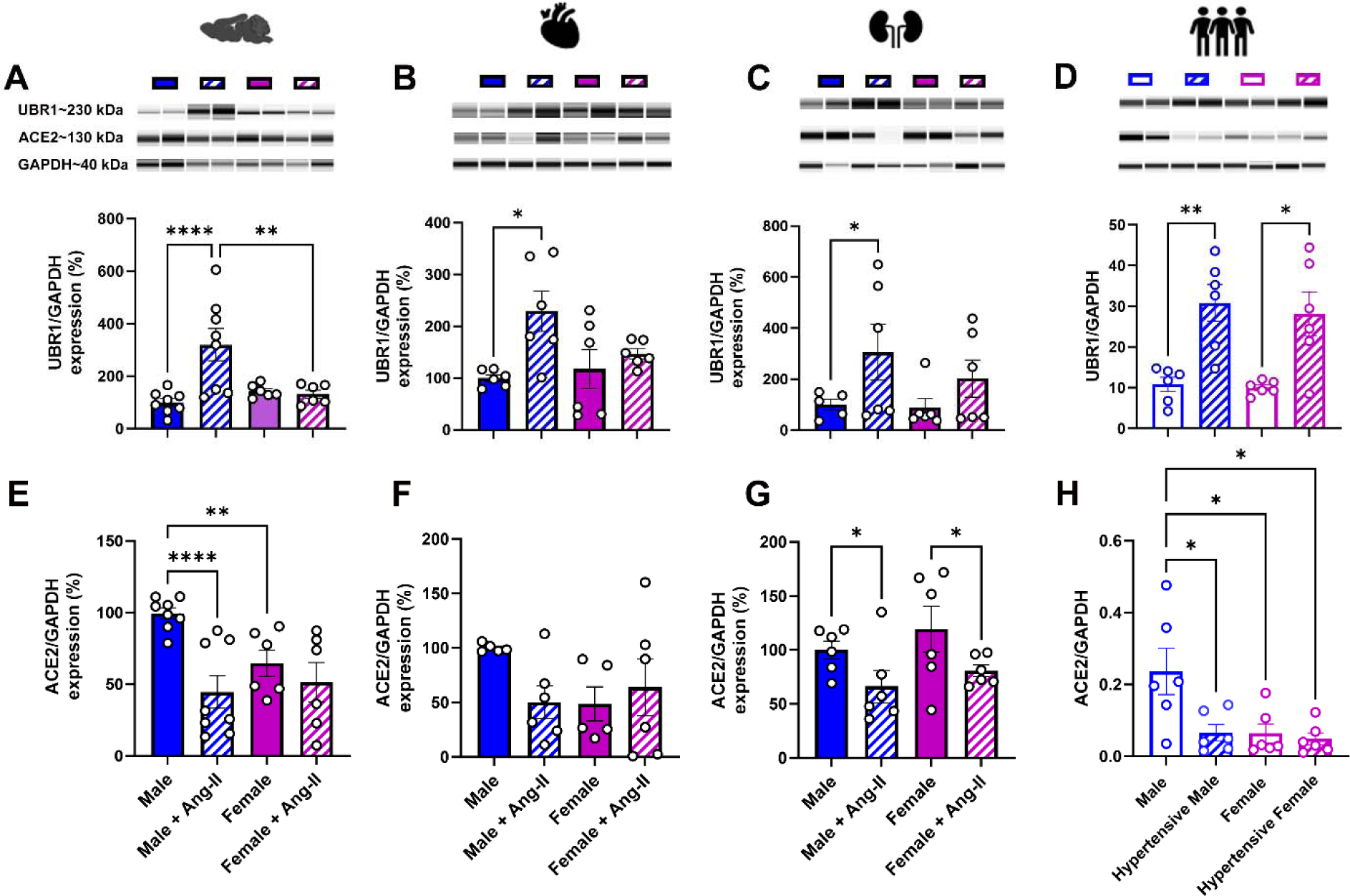
UBR1 and ACE2 expression are differentially regulated by sex hormones in Ang-II-mediated hypertension. Expression of UBR1 (A-C) and ACE2 (E-G) was assessed in the mouse hypothalamus, heart, and kidney (n= 6-8/group) after 4 weeks of Ang-II infusion and in human cardiac left ventricle samples (D,H). Representative immunoassays for UBR1, ACE2 and GAPDH samples from Caucasian and Black donors (n=6-7/group) (D,H). Data are shown as mean ±SEM. Statistical significance: Two-way analysis of variance (ANOVA): ****P<0.0001, **P<0.01 and *P<0.05. Abbreviations: NT, normotensive; HT, hypertensive.

### UBR1 contributes to Ang-II-mediated ACE2 downregulation

To assess causality between UBR1 upregulation and ACE2 downregulation, primary human aortic endothelial cells (HAEC) were transfected with a UBR1 siRNA which led to a dose-dependent upregulation of ACE2 (Figure 3A). Pre-treatment with UBR1 siRNA produced a significant knockdown of UBR1 expression (***P<0.001), associated with a parallel increase in ACE2 levels (**P<0.01), demonstrating that UBR1 constitutively regulates ACE2 expression. As observed in mice (Figure 2), Ang-II treatment of HAEC led to a significant upregulation of UBR1, associated with a reduction of ACE2 protein levels (Figure 3B; ***P<0.001). Immunolabeling (Figure 3C) showed that in baseline conditions ACE2 is expressed on the cell surface of HAEC while UBR1 is located throughout the cell surface and cytoplasm. Upon treatment with Ang-II, ACE2 was internalized in the cytoplasm but also in the nucleus where it strongly co-localized with UBR1. Importantly, silencing of UBR1 prevented not only prevented the Ang-II mediated upregulation of this E3 ligase but also resulted in an increase in ACE2 levels (Figure 3B; ***P<0.001). These data suggest that UBR1 is critical for Ang-II-mediated downregulation of ACE2. Since UBR2 has been shown to functionally overlap with UBR1, the possibility that UBR2 could also ubiquitinate ACE2 was further investigated. As suspected, transfection with UBR2 in HEK293T cells reduced ACE2 expression and activity (Figure S2 in data supplement) but this effect was independent of Ang-II. Together, these data confirm that UBR1 is a newly identified mediator of ACE2 ubiquitination.

**Figure 3:**
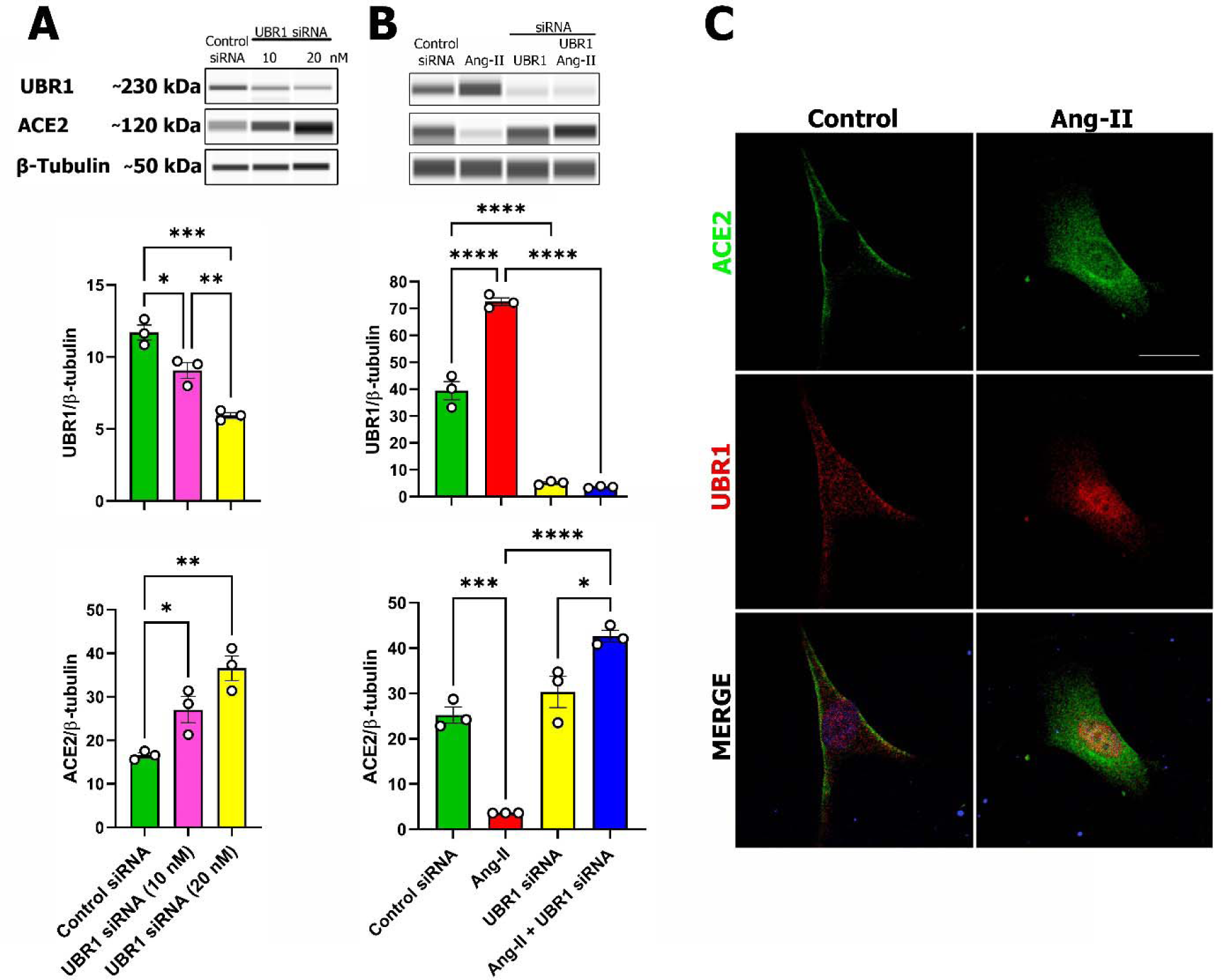
UBR1 contributes to Ang-II mediated ACE2 degradation. Human aortic endothelial cells (HAEC) were pre-treated with control (scramble) or UBR1 siRNA for 6 hours before exposure to Ang-II (100 nM) for 4 hours. Capillary Western Blot showing a UBR1 siRNA dose-response increase in UBR1 protein level and decrease in ACE2 levels **(A).** Capillary Western Blot showing a restoration of ACE2 levels following the knockdown of HAEC with UBR1 siRNA **(B).** GFP-labeled ACE2 (Green) localizes at the plasma membrane in control cells and internalizes to the cytoplasm after Ang-II treatment. Texas red-labelled UBR1 is present throughout the cell and colocalizes with ACE2 mostly in the nucleus (DAPI, blue) following Ang-II treatment **(C)**. Data are shown as mean ±SEM (n=3, in triplicate). Statistical significance: One-way analysis of variance (ANOVA) followed by Bonferroni test for multiple comparisons, *P<0.05, **P<0.01,***P<0.001, ****P<0.0001

### Role of sex hormones in UBR1 expression

To further address the role of sex hormones on ACE2 ubiquitination by UBR1, we expanded our experiments to HAEC and smooth muscle cells (SMC) to test for possible cell specificity. Similar to the lack of UBR1 upregulation in female mice (Figure 2), 17β-estradiol treatment failed to increase UBR1 expression in HAEC (Figure 4A,B) or SMC (Figure 4A,C), in the presence or absence of Ang-II. While 17β-estradiol did not affect ACE2 expression (Figure 4A,E) in HAEC, blockade of either alpha or beta estrogen receptors (using MPP and PHTPP, respectively) prevented the Ang-II mediated upregulation of UBR1 and concomitant downregulation of ACE2 (Figure 4A,B,D) in HAEC, suggesting that 17β-estradiol might have a synergistic action with Ang-II on UBR1 expression. Unlike 17β-estradiol, DHT alone increased UBR1 expression in SMC, but not in HAEC (Figure 4A-C). In both cells, this was associated with reduced ACE2 expression independently of Ang-II (Figure 4A,D,E). While androgen receptor blocker flutamide was ineffective, likely due to incomplete prevention of DNA binding, these responses could be prevented by the androgen receptor DNA binding domain inhibitor (DBDi). Together, these data show that male sex hormones appear to upregulate UBR1 and simultaneously reduce ACE2 in a cell specific manner while 17β-estradiol has no direct impact on UBR1.

**Figure 4:**
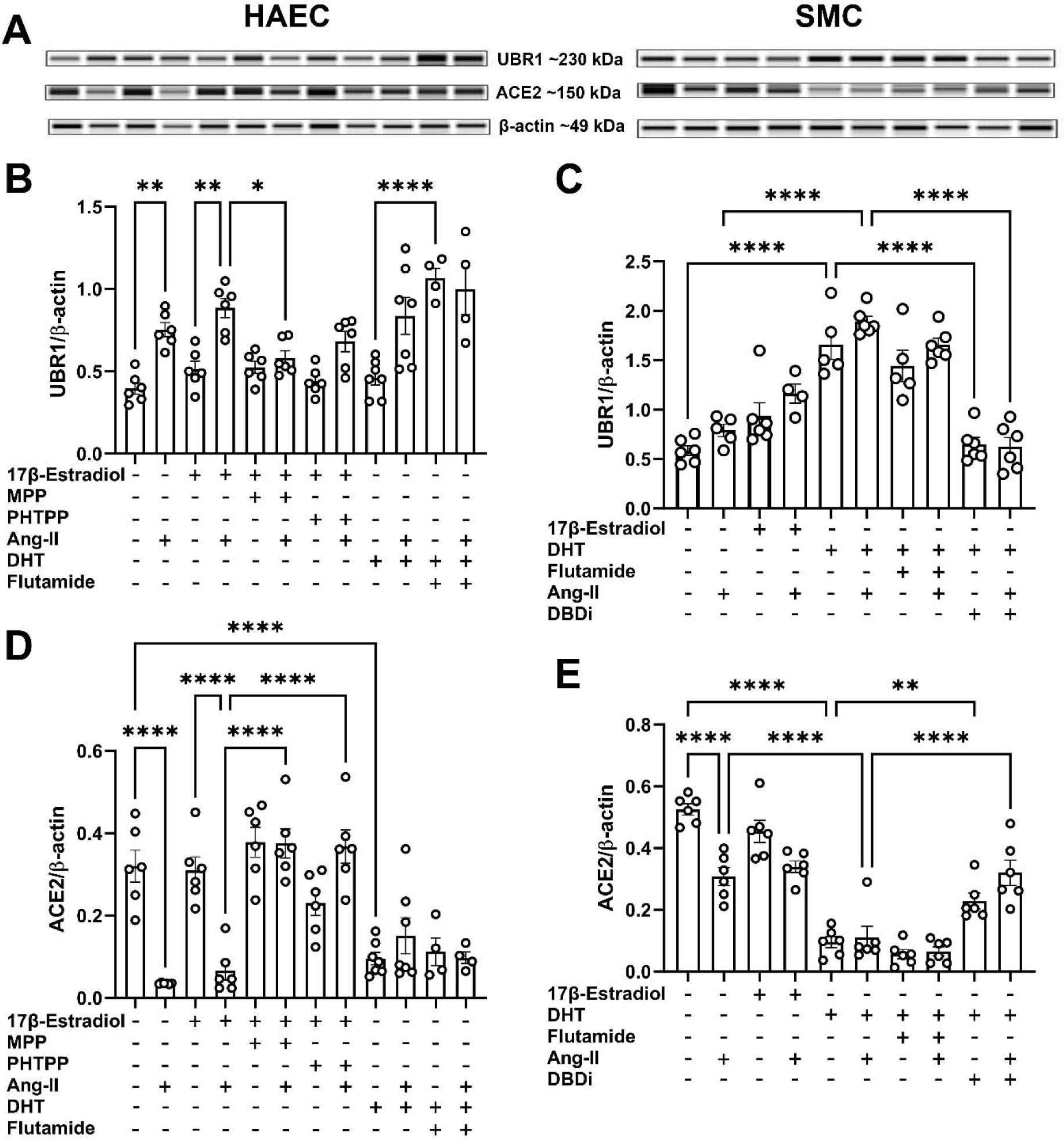
Sex hormones regulation of UBR1. Protein expression of UBR1 and ACE2 in human aortic endothelial cells (**A,B**) and smooth muscle cells (**C,D**). Expression was normalized to β-actin and β-tubulin housekeeping proteins. MPP (Methylpiperidinopyrazole): selective estrogen receptor alpha blocker; PHTPP: selective estrogen receptor β antagonist; DHT: dihydrotestosterone; DBDi: DNA binding domain inhibitor. One way ANOVA followed by Bonferroni test for multiple comparisons, *P<0.05, **P<0.01,***P<0.001, ****P<0.0001. N=4-6/group.

### UBR1 knockdown restores ACE2 levels in hypertensive mice

Based on our observations that UBR1 is only upregulated in males, we proceeded to investigate the role of UBR1 in ACE2 ubiquitination and hypertension. C57Bl6/J males (12-week-old, n=5/group) were chronically infused with Ang-II for 4 weeks while treated centrally or peripherally to achieve UBR1 knockdown (Figure 5A). UBR1 silencing following the establishment of hypertension had no immediate effect when administered centrally but was associated with an increase in BP in the first week of treatment, when infused peripherally (Figure 5B). A transient reduction in BP occurred in the central UBR1 siRNA group after 3 weeks of UBR1 knockdown (Figure 5B,C), most notably in the resting phase of the nychthemeral cycle. In both peripheral and central UBR1 siRNA groups, Ang-II-induced upregulation of UBR1 was blunted (Figure 6A), and this was paralleled with a restoration of ACE2 protein levels in the hypothalamus (Figure 6B). Interestingly, the Ang-II mediated upregulation of UBR1 was associated with an increase in another E3 ligase expression, Nedd4-2 (Figure 6C), its inactivated form, phosphorylated Nedd4-2 (Figure 6D,E) and the kinase responsible for its inactivation, serum and glucocorticoid-regulated kinase 1 (SGK1, Figure 6F). The activation of Nedd4-2 was evidenced by a decrease in phosphorylated Nedd4-2 relative to total Nedd4-2 in UBR1 siRNA groups (Figure 6 C-E). Based on these observations, mice were transfected icv with a lentivirus encoding a scrambled or Nedd4-2 shRNA (Figure S3 in data supplement). While baseline BP was not affected by the treatment, a blunted hypertension was noted early on post Ang-II infusion. However, this reduction of hypertension did not persist and was gone by the third week of Ang-II treatment. To further understand the mechanism of Nedd4-2 activation, we measured the protein expression of SGK1, the kinase responsible for Nedd4-2 phosphorylation. Indeed, SGK1 protein levels were also reduced in UBR1 siRNA treated groups, supporting the overall activation of Nedd4-2 in UBR1 knockdown mice (Figure 6F).

**Figure 5:**
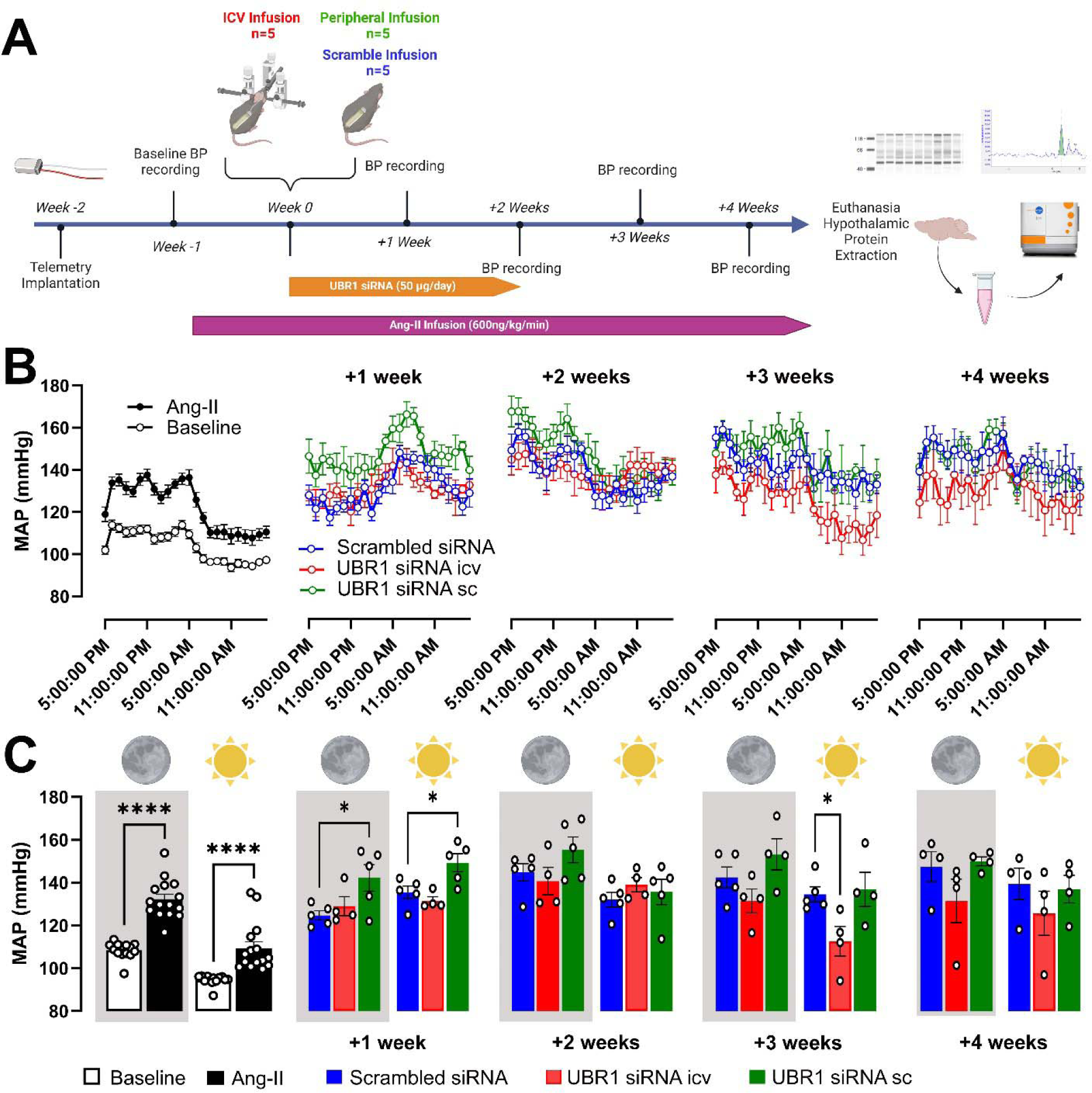
UBR1 siRNA Infusion in Hypertensive Mice. Experimental timeline of UBR1 siRNA infusion (**A**). Weekly MAP recording in mice infused with Ang-II combined with ICV or subcutaneous UBR1 siRNA (**B**). Summary data showing the average MAP per week in different groups (**C**). One-way ANOVA. *P<0.05, ****P<0.0001, n=4-15/gro-up.

**Figure 6:**
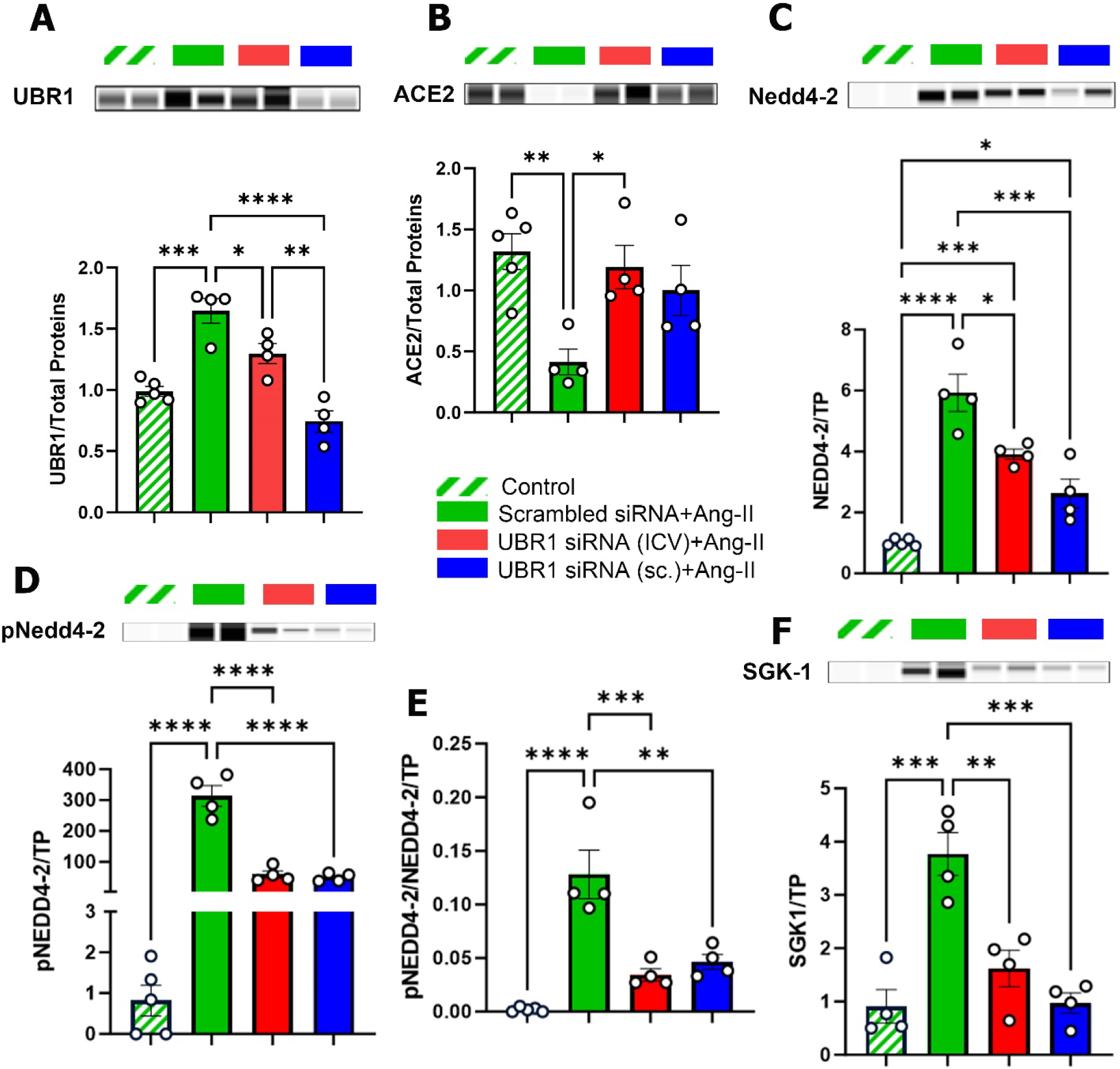
Protein Expression in UBR1 siRNA-Infused Hypertensive Mice. Capillary Western showing protein levels in the hypothalamus for UBR1 (**A**, ∼230 kDa), ACE2 (**B**, ∼140 kDa), Nedd4-2 (**C**, ∼130 kDa), pNedd4-2 (**D**, ∼130 kDa), pNedd4-2 to Nedd4-2 ratio (**E**), and SGK1 (**F**, ∼49 kDa) in various treatment groups. TP: total protein. One-way analysis of variance (ANOVA) followed by Bonferroni test for multiple comparisons, ****P<0.0001; ***P<0.001; **P<0.001; *P<0.05, n=4-5/group.

## Discussion

Although ACE2 was discovered in 2000, it took almost 15 years for the first ACE2 ubiquitination data to be reported.^2^ MDM2 was then identified as the first E3 ligase targeting ACE2^10^ and our group recently reported the role of Nedd4-2 in ACE2 ubiquitination.^3^ Using a proteomics discovery approach, followed by a multilayer’s validation process, we now report the identification of UBR1 as a the first E3 ligase from the N-end rule pathway to ubiquitinate ACE2. Moreover, our data show that UBR1 is differentially expressed during hypertension and regulated in a sex-dependent manner. Our group previously reported that upon Ang-II treatment, ACE2 moves from the cell surface to the lysosomes, followed by degradation in lysosomes, leading to a loss of ACE2 expression and compensatory activity in diseases such as hypertension.^2^ This process involving ubiquitination is dependent on AT_1_R, and was also observed in the case of S1 spike protein (component of SARS-CoV-2 envelope)-induced ACE2 internalization.^4^ Ang-II activation of E3 ligases has long been recognized, notably for Nedd4-2 and Cullin3.^7^ Expression of ACE2 has also been shown to modulate the levels of E3 ligases, for example Smurf2 which is upregulated in the absence of ACE2.^26^ The COVID-19 pandemic provided a strong impetus to understand the mechanisms involved in ACE2 post-translational regulation and several new ubiquitases have been suggested,^1,27^ although most of them have not been associated with cardiovascular regulation. The exception is our recent identification of Nedd4-2 as an E3 ligase targeting ACE2 in pre-clinical and clinical hypertension.^3^ In this study we identified and validated UBR1 as a new E3 ligase targeting ACE2. Additionally, we show a synergistic interaction between Nedd4-2 and UBR1.

We observed that experimental and clinical hypertension is associated with an upregulation of the E3 ligase UBR1, notably in males. In addition, although some cell specificity was noted, all major tissues involved in BP regulation (*i.e.* brain, heart and kidney) exhibited a similar sex difference. The role of UBR1 in ACE2 ubiquitination was further confirmed in loss of function experiments showing that upon silencing UBR1, Ang-II is no longer able to induce ACE2 internalization and degradation. UBR1 is an E3 ligase part of the N-end rule pathway, that binds and destabilizes N-terminal residues, such as arginine, lysine, histidine, and bulky hydrophobic residues, promoting polyubiquitination and the eventual degradation of the substrate. Although UBR1 was primarily reported to modulate proteasomal degradation,^28^ it also plays a role in the quality control process, independent of the N-end rule pathway.^29^ Interestingly, UBR1 also modulates proteolysis of the regulators of G-protein signaling, RGS4 and RGS5, and differentially controls G-protein-coupled receptor signaling under normoxic/hypoxic conditions.^30^ Although mutations of the UBR1 gene have been associated with Johanson-Blizzard syndrome, a rare genetic disorder that affects multiple organs,^28^ to date, there is little information about UBR1 suggesting it plays a role in cardiovascular diseases. Previous work reported that pericytes infected by the Japanese encephalitis virus could lead to a compromised blood-brain barrier mediated via UBR1.^31^ This is relevant since experimental models of hypertension with increased Ang-II expression increase blood-brain barrier permeability.^32^ In addition, ACE2 has also been reported to contribute to blood-brain barrier homeostasis.^33^ Although not explored in our study, these observations could support a role for UBR1 in blood brain barrier leakage in cardiovascular conditions. Our data show that UBR1 upregulation in hypertension was not restricted to the brain but also apparent in the heart and kidney. Moreover, silencing of UBR1 in endothelial cells not only prevented Ang-II-mediated downregulation of ACE2 but was associated with an up-regulation of ACE2. Therefore, it is conceivable that Ang-II-mediated upregulation of UBR1 in endothelial cells promotes ACE2 degradation, which in the brain could contribute to blood-brain barrier dysfunction and the establishment of neurogenic hypertension. Downregulation of ACE2 from endothelial cells in the heart and kidneys could also contribute to cardiovascular disease progression through multiple mechanisms, including impaired endothelial function.

Another finding from our study is the major role played by sex hormones on ACE2 expression and its ubiquitination partners. Clinical and pre-clinical studies have reported opposite effects of sex hormones on ACE2 activity and expression.^34–38^ In line with our observations, ACE2 expression is thought to be lower in females than males in multiple cell types, tissues and species,^34,35,38^ while gonadectomy reverses these findings.^37^ However, unlike our findings, exposure to testosterone led to increased ACE2 expression in pulmonary cells from both males and females while 17β-estradiol downregulated ACE2.^34,36^ Intriguingly, expression levels of fascin-1, an actin bundling protein that protects ACE2 against Ang-II downregulation,^39^ inversely correlates with estrogen receptor alpha expression,^40,41^ indicating that multiple steps in ACE2 internalization and degradation can be modulated by sex hormones. In our study, DHT upregulated UBR1 expression in SMC but not in endothelial cells, and this was associated with a reduction of ACE2. This sex specificity is further supported by a previous report showing that UBR1 promotes the degradation of the androgen receptor.^42^ To the best of our knowledge, the present study is the first to show the impact of sex hormones on ACE2 ubiquitination.

Our validation experiments in cells clearly show the co-dependence between Ang-II and UBR1, with Ang-II promoting UBR1 upregulation and its inability to induce ACE2 downregulation when UBR1 has been silenced. Yet, some key questions remain unanswered. Although our data show a massive upregulation of UBR1 in hypertensive experimental conditions associated with increased Ang-II levels, as well as clinical hypertension, we failed to see a major effect of UBR1 silencing on the development of hypertension. This could be partially explained by the concomitant upregulation of active Nedd4-2 and possibly other pro-hypertensive E3 ligases which could have mitigated the impact of UBR1 knockdown by inducing pro-hypertensive pathways. Although associated with developmental abnormalities and cognitive deficits,^43^ complete deletion of UBR1 in mice is viable further supporting compensation by other UBR1 homologs, for example UBR2, or other E3 ligases. While knockdown of UBR1 had only a modest impact on hypertension, it was sufficient to restore ACE2 expression in the hypothalamus. UBR1 knockdown induced a compensatory positive feedback triggering Nedd4-2 activation which we speculate prevented the reduction of hypertension. A similar lack of efficiency was also observed when Nedd4-2 knockdown in the brain was initiated prior to the development of hypertension (Figure S3 in data supplement), where a transient drop in BP was observed followed by an increase in BP.

Interestingly, the type of ubiquitination driven by UBR1 is also thought to regulate protein function, and as such, it has been involved in trafficking, activation of signaling pathways, as well as synaptic plasticity of excitatory neurons.^44,45^

Another unresolved question is the trafficking of ACE2 to the nucleus. Other members of the RAS, notably AT_1_R, have previously been identified in the nucleus, which forms the basis for the existence of an intracellular RAS.^46^ To our knowledge, there is no previous report of ACE2 being identified in this organelle and further research is needed to confirm this observation and determine if it is associated with a regulatory function.

In summary, the present study identifies a novel E3 ligase involved in ACE2 ubiquitination, taking place in various organs in experimental and clinical hypertension and potentially capable of modulating ACE2 compensatory activity during the development of cardiovascular diseases. Sex differences were further shown to play a critical role in the expression levels of ACE2 and its ubiquitination. We propose that ACE2 ubiquitination is an important mechanism leading to Ang-II-mediated internalization of this enzyme and the loss of its compensatory activity. Inhibition of UBR1 could be a new therapeutic strategy to prevent ACE2 ubiquitination in hypertension.

## Novelty and Significance

### What is known?

- ACE2 undergoes Ang-II-induced internalization via AT_1_R, followed by ubiquitination and lysosomal degradation during hypertension.
- Murine double minute 2 (MDM2) and S-Phase Kinase Associated Protein 2 (Skp2) and Nedd4-2 are E3 ligases that promote ACE2 ubiquitination.
- Estrogens and testosterone affect the expression and activity of RAS components, contributing to sex differences in cardiovascular diseases.

### What new information does this article contribute?

- UBR1 is a new E3 ligase involved in ACE2 ubiquitination in hypertension, especially in males. The increase in UBR1 in hypertensive conditions is differentially regulated by sex hormones.
- Ang-II-mediated ACE2 internalization, ubiquitination and degradation is taking place within various tissues, including brain, heart and kidney in mice and is clinically relevant to hypertensive conditions, as indicated by reduced ACE2 levels in cardiac samples from hypertensive patients.
- UBR1 works synergistically with Nedd4-2, another E3 ligase, to regulate ACE2 and BP levels.
- UBR1 could be a new therapeutic target to preserve ACE2 compensatory activity in hypertension.
- This work provides new insights into the regulation of ACE2 and highlights pathways that could be targeted for BP regulation.

## Supporting information

Main manuscript

## Data Availability

Data will be made available at the time of publication.

## Acknowledgments

The authors would like to thank the Wisconsin Donor Network and Versiti Blood Center of Wisconsin organ procurement and Duke University teams for obtaining tissue from rejected donor organs.

## Authors contributions

M. Elgazzaz, N. Lakkappa, C. Berdasco, X. Yue, F. Mauvais-Jarvis, X. Yue, C.M. Filipeanu and E. Lazartigues designed experiments. M. Elgazzaz, N. Lakkappa, C. Berdasco, J.J. Guidry, S. Sriramula, J. Xu, U.P. Mohan, L. Restivo, A. Martinez, A. Scarborough, X. Yue, and H. Daoud performed the experiments. A.M. Beyer, M.A. Mendiola and D.E. Bowles contributed de-identified patient samples. M. Elgazzaz, N. Lakkappa, X. Yue, C.M. Filipeanu and E. Lazartigues wrote the manuscript. All authors contributed to manuscript revision.

---

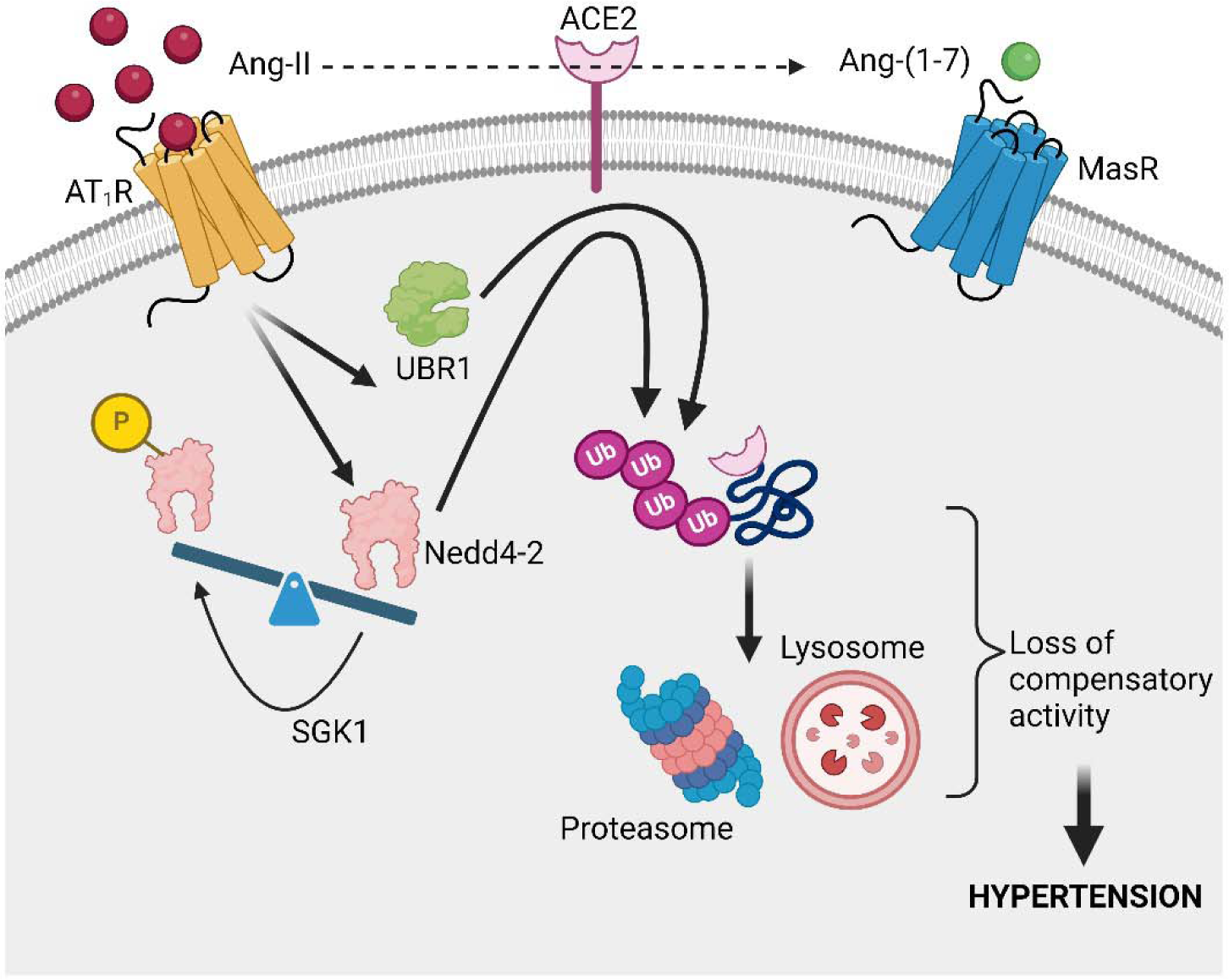
Graphical abstract: Ang-II promotes ACE2 ubiquitination and hypertension. Ang-II binding to AT_1_R is associated with upregulation of the E3 ligases UBR1 and Nedd4-2, mostly in males. UBR1 and enhanced active Nedd4-2 both contribute to ACE2 polyubiquitination eventually leading to ACE2 degradation, the loss of compensatory activity and hypertension.

## Nonstandard Abbreviation and Acronyms

ACE2: Angiotensin Converting Enzyme 2
Ang-II: Angiotensin II
Ang-(1-7): Angiotensin (1-7)
ANOVA: Analysis of variance
AT_1_R: Ang-II type 1 receptor
BP: Blood pressure
DHT: dihydrotestosterone
DBDi: androgen receptor (AR) DNA-binding domain inhibitor
E1: Ubiquitin-activating enzymes
E2: Ubiquitin-conjugating enzymes
E3: Ubiquitin protein ligases
ENaC: Kidney epithelial Na^+^ channels
HAEC: Human aortic endothelial cells
LC-MS: Liquid chromatography–mass spectrometry
MAP: Mean arterial pressure
MDM2: Murine double minute 2
MPP: Methylpiperidinopyrazole
Nedd4-2: Neural Precursor Cell Expressed Developmentally Down-Regulated Protein 4
PHTPP: 4-[2-PHENYL-5,7-BIS(TRIFLUOROMETHYL)PYRAZOLO[1,5-A]PYRIMIDIN-3-YL]PHENOL
RAS: Renin-angiotensin system
Skp2: S-Phase Kinase Associated Protein 2
siRNA: Small interfering RNA
SMC: Smooth muscle cell
SGK1: Serum/Glucocorticoid-Regulated Kinase 1
UBR1: Ubiquitin Protein Ligase E3 Component N-Recognin

