## Supplementary material for "UBR1 Promotes Sex-Dependent ACE2 Ubiquitination in Hypertension": Main manuscript

Running title: ACE2 ubiquitination in hypertension

Number of words: 2722

^#^Correspondence to:

Dr. Eric Lazartigues, PhD; Louisiana State University Health, School of Medicine, Cardiovascular Center of Excellence, 533 Bolivar Street, New Orleans, LA 70112.

**Methods**

The authors declare that all supporting data are available within the article and in the Data Supplement.

Experiments were conducted in adult C57BL6/J mice (8–12 weeks old, 20-25 g; Jackson Laboratory, Bar Harbor, ME) from both sexes. Mice were housed in a temperature (~25 ºC) and humidity-controlled facility under a reversed 12-hour dark/light cycle, fed standard mouse chow (Envigo, iOS Teklab Extruded Rodent Diet 2019S, Huntingdon, UK) and water ad libitum. All procedures conformed to the National Institutes of Health Guide for the Care and Use of Laboratory Animals and were approved by the Louisiana State University Health Sciences Center (#3540), and the Southeast Veterans Healthcare System (#620) Institutional Animal Care and Use Committees in accordance with the ‘Principles of Laboratory Animal Care by the National Society for Medical research and the Guide for the Care and Use of Laboratory Animals’ (National Institutes of Health Publication No. 86-23, revised 1996). Cardiac samples from patients were obtained from The Medical College of Wisconsin (IRB #PRO00010828) and Duke University (IRB #PRO00005621) from organs not suitable for transplant. Informed consents were obtained prior to experiments and all procedures conform to the principles outlined in the Declaration of Helsinki.

*Telemetry probes implantation*

Before surgery, mice were anesthetized with isoflurane (2%) in an oxygen flow (1 L/minute) and placed on a heating pad to maintain body temperature around 37.5°C. Mice were implanted with telemetry probes (PA-C10 or HD-X10; DSI, St. Paul, MN) for conscious blood pressure (BP) recording, as reported previously.^1^ Following recovery, baseline BP was recorded weekly for 24 hours in all groups.

*Ang-II infusion model*

Ang-II (600 ng/kg/min; SigmaAldrich, St Louis, MO, A9525) or vehicle (0.9% saline) infusion was performed by implanting subcutaneous osmotic mini pumps (Alzet Model 1004; Durect Corporation, Minneapolis, MN) for a period of 4 weeks.

After completion of recording, the mice were euthanized by decapitation followed by blood collection. Brain, heart, and kidneys were collected from each group (6 mice/group) and were snap-frozen in liquid nitrogen for further analysis.

*Proteomics / LCMS*

Discovery-based Proteomics using Tandem Mass Tags (TMTpro) and Liquid Chromatography Mass Spectrometry (LC-MS): Samples were prepared for discovery-based quantitative proteomic analysis by the addition of SDS to 1% and sonicated until completely homogenous. The protein concentration was determined using BCA protein assay kit (Pierce, Thermo Fisher Scientific) using an eight-point standard curve. Protein samples were prepared for trypsin digestion by reducing the cysteines with tris(2-carboxyethyl) phosphine (TCEP) followed by alkylation with Iodoacetamide (IAA). After Chloroform-Methanol precipitation, each protein pellet was digested with 1 µg trypsin overnight at 37 °C. The digested product was labeled using 2 - TMTpro 16 plex Reagents Sets (Thermo Fisher Scientific Pierce), utilizing the 126 isotopologue as the common pooled internal control, according to the manufacturer’s protocol.

An equal amount of each TMTpro-labelled sample was pooled together in a single tube and SepPak purified (Waters, Ireland) using acidic reverse phase conditions to remove unreacted TMTpro and quenched-TMTpro molecules. After drying to completion, an off-line fractionation step was employed to reduce the complexity of the sample using basic pH reverse phase chromatography (Dionex U3000, Thermo Fisher Scientific). Each fraction was subjected to a 95-min chromatographic method employing a gradient from 2-25% ACN in 0.1% formic acid (FA) (ACN/FA) over the course of 65 min, a gradient to 50% ACN/FA for an additional 10 min, a step to 90% ACN/FA for 5 min, and a 15-min re-equilibration into 2% ACN/FA. Chromatography was carried out in a “trap-and-load” format using an EASY-Spray source (Thermo Fisher Scientific); trap column C18 PepMap 100, 5 µm, 100 A and the separation column was an EASY-Spray PepMap RSLC C18 2 µm, 100A, 75 µm x 25 cm (Thermo Fisher Scientific Dionex, Sunnyvale, CA). The entire run was at a flow rate of 0.3 µl/min. Electrospray was achieved at 1.8 kV. TMTpro data acquisition utilized an MS3 approach for data collection. The Protein FASTA database was Mus musculus, SwissProt tax ID=10090, version 2017-10-25 and contained 25,097 sequences. Static modifications included TMTpro reagents on lysine and N-terminus (+304.207), carbamidomethyl on cysteines (+57.021), dynamic phosphorylation of Serine, Threonine and Tyrosine (+79.966 Da), and dynamic modification of oxidation of methionine (+15.9949). The mass spectrometry proteomics data has been deposited to the ProteomeXchange Consortium via the PRIDE partner repository with the dataset identifier PXD (PXD027183).

*Cell culture and maintenance*

Primary Human Aortic Endothelial Cells (HAEC; PCS­100­ 011, ATCC, USA) were obtained from ATCC^®^ (Manassas, VA) and cultured in Endothelial Cell Growth Kit­VEGF (ATCC® PCS­100­041). Human Embryonic Kidney 293T cells (ATCC^®^ CRL-3216™) were cultured in Dulbecco’s Modified Eagle’s Medium (DMEM) (ATCC 30-2002), 10% Fetal Bovine Serum (heat inactivated) (ATCC 30-2020), 2 mM L-glutamine (ATCC 30-2214) as per the manufacturer’s instructions. Upon reaching ~90% confluency, the cells were trypsinized and seeded in six‐well plates at a density of 3 × 10^5^ cells per well in a culture medium (2 mL) for 24 h. The cells were serum-starved 24 h before each experiment. Primary Human Aortic Smooth Muscle Cells (HASMC, PCS-100-012, ATCC, USA) were seeded and maintained according to the manufacturer protocol using Vascular Smooth Muscle Cell Growth Kit (PCS-100-042, ATCC). Briefly, growth kit contents were transferred into a vascular cell basal media bottle (PCS-100-030, ATCC). Cells were added to the complete media after thawing. Cells were passaged after 80% confluence. For investigating the sex-driven effects on UBR1, we used HAEC and HASMC. At passage 2, cells were seeded into a 12 well plate and left for 24 h to allow adherence to the plate bottom. Cells were either treated with vehicle; Ang-II (100 nM, 4h); 17β Estradiol (10 nM); MPP (1 µM); PHTPP(1 µM); DHT (10 nM); Flutamide (10 µM); DBDi (10 µM) or a combination. Following treatment, cells were harvested for protein extraction using lysis buffer for protein quantification studies.

*siRNA Transfection*

The siRNA and Lipofectamine RNAiMAX Transfection Reagent complex were prepared at different ratios for optimum standardization (10, 20 and 30 nM of UBR1‐targeted siRNA (ID: 129855, ThermoFisher Scientific) with 1, 3 or 12 µl of RNAIMAX transfection reagent accordingly) in Opti-MEM Reduced Serum Media. The complexes were incubated for 5 min at room temperature, added to each well in 6 well plates containing endothelial cells and incubated in an antibody-free medium. After 6 h, the medium was replaced, and the endothelial cells were allowed to grow for 24 h. The cells were then trypsinized and harvested for quantification of relative UBR1, expression by capillary Western assay. The siRNA concentration of 20 nM and 3 µl RNAIMAX reagent were narrowed down for further experiments. The cells were transfected with 20 nM UBR1 siRNA, or a scrambled control siRNA. After 24 h, the cells were stimulated with Ang-II (100 nM) for 4 h and harvested for quantification of UBR1, β-tubulin and ACE2 by capillary western analysis (Protein Simple). A similar protocol was used in another set of cells for immunohistochemistry (IHC).

*Immunocytochemistry*

HEK293T and primary human aortic endothelial cells (HAEC) were seeded on poly-L-lysine-coated cover slips placed in 6 well plates. After 24 h, the cells were rinsed with 1X PBS, and fixed with 4% PFA at room temperature (RT) for 15 min. The cells were washed twice with 1X glycine PBS (250 mM Glycine in 1X PBS) at RT for 15 min. Further, the cells were treated with permeabilization buffer (20 µL 100X triton in 10 mL 1X PBS) at RT for 15 min then washed with 1X PBS at RT for 15 min. To prevent non-specific binding of antibodies or other reagents to the tissue, blocking was carried out using 10% Normal horse serum + 0.3%Triton 100X at RT for 30 min. The blocking was replaced with Primary antibody (ACE2 or UBR1) diluted to 1:500 in 2% normal horse serum + 0.3% Triton 100X at 4 °C, overnight. The cells were further washed 3 times with PBS at RT for 10 min. The cells were treated with respective secondary antibodies at 1:1000 dilution in 2% normal horse serum + 0.3% Triton 100X at RT for 1 h. Followed by washing in PBS for 3 times at RT for 10 min and the cover slips were mounted on slides using mounting media containing DAPI (Fluoromount-G™ Mounting Medium, 00-4959-52, Invitrogen™). The imaging was done using a confocal microscope (Eclipse Ti2, Nikon, Japan) and images were processed using NIS-Element’s software.

*Capillary Western*

Total proteins from tissues were extracted using a RIPA lysis buffer containing protease (Pierce^TM^ RIPA buffer, 89901, Thermo Fisher Scientific, IL and Protease inhibitor cocktail tablets, 04693159001, Roche diagnostic, Germany) while cells were extracted using a cell lysis buffer (R&D systems, 895347) containing proteases and phosphatases inhibitors and quantified using BCA protein assay kit (Pierce, Thermo Fisher Scientific). Equal amounts of proteins were loaded and analyzed on a capillary-based immunoassay platform (Wes™, ProteinSimple, San Jose, CA) as per the manufacturer's instructions using a 12–240 kDa Separation Module (Bio-Techne R&D Systems, SM-W004). The samples were diluted to 0.5 µg/ml concentration in the sample buffer (10X Sample Buffer-2 was diluted to 100x from the Separation Module (Bio-Techne R&D Systems 042-195)), incubated with a Fluorescent Master Mix (Protein Simple PS-ST01EZ) and heated at 95 °C for 5 min. The Biotinylated Protein Ladder (Protein Simple PS-ST01EZ), samples, blocking reagent (Protein Simple 042-203), primary antibodies: ACE2 (SC #390581, Santa Cruz Biotechnology), GAPDH (ab #9483, Abcam), β-Tubulin (ab #21058, Abcam), UBR1 (SC #515753, Santa Cruz Biotechnology), and UBR1 (ab138267, Abcam), pNedd4-2 (PA5-104619, Invitrogen ), Nedd4-2 (2740S, Cell Signaling Technology), SGK-1 (GTX107750, GeneTex) were used at concentration of 1:10 and HRP-conjugated secondary antibodies (042-205, 042-206 and 043-522-2) ready to use from ProteinSimple™, and the luminol-peroxide mixture (ProteinSimple™ 043-311 and 043-379) as chemiluminescent substrate were pipetted into the plate. The plate and cartridges were loaded into the Wes with the following setting: stacking and separation at 475 V for 30 min; blocking reagent for 5 min; primary and secondary antibody both for 30 min. Compass software was used to process and analyze the results.

UBR1 siRNA Infusions

UBR1 siRNA (#4457310, Assay ID; s75706, SilencerSelect, Thermofisher Scientific, USA) was used to knockdown UBR1 expression in mice. Ten weeks old male mice were implanted with radiotelemetry probes and baseline BP was recorded following 10 days recovery. Mice were then implanted with subcutaneous osmotic pumps filled with Ang-II (600 ng/Kg/min, Alzet Model 1004; Durect Corporation, Minneapolis, MN) to induce hypertension. One week after Ang-II infusion, mice were subdivided into 3 groups and chronically infused with one of the following treatments (50 µg/day/mouse): 1) ICV UBR1 siRNA ; 2) subcutaneous peripheral (sc.) UBR1 siRNA; 3) scrambled siRNA. Treatments were delivered using mini osmotic pumps for 2 weeks (Alzet Model 1002; Durect Corporation, Minneapolis, MN). For ICV group, the skull was stabilized on a stereotaxic apparatus (Stoelting, USA). A hole in the skull was made and a canula was inserted (coordinates; AP: 0.4 mm, ML: ±1 mm, DV: -3 mm) and connected to a sc. mini osmotic pump via a catheter for ICV drug delivery. For the two other groups, osmotic pumps were inserted underneath the skin. Hemodynamic parameters were recorded once a week for 4 weeks after siRNA infusion. To maintain Ang-II infusion for the total 5 weeks of BP recording, Ang-II osmotic pumps were replaced after 4 weeks of infusion with another 2-week pump (600 ng/Kg/min, Alzet Model 1002, Durect, USA).

*Nedd4-2 ShRNA Virus Injection*

C57BL/6J male mice (10/group) were anesthetized using 2.5% isoflurane. The skull was stabilized on a stereotaxic apparatus (Stoelting, USA). The mice received bilateral injection in the PVN (coordinates; AP: 0 mm, ML: ±0.25 mm, DV: -4.7 mm) of lentivirus containing either Nedd4-2 or scrambled ShRNA (100 nl, 1.7X107 TU/ml, GeneCopoeia, USA) using Pneumatic PicoPump (WPI, USA). After 3 weeks of virus injections, mice were implanted with sc Ang-II osmotic pumps (600 ng/Kg/min, Alzet Model 1004; Durect Corporation, Minneapolis, MN). BP was recorded using telemetry one week after for 24 hr for 4 consecutive weeks.

*ACE2 activity*

HEK293T cells were processed for ACE2 activity. Briefly, the cells were suspended in 0.5% Triton X-100 in ACE2 reaction buffer containing 1 M NaCl, 0.5 mM ZnCl2, 75 mM Tris·HCl and 100 μM Mca-YVADAPK(Dnp). Fluorescence emission at 405 nm, after excitation at 320 nm, was measured and the slope of fluorescence development between 10 and 120 min of incubation was calculated. Data are presented as fluorescence units (amount of substrate converted to fluorescent product) per minute normalized for protein content (FU/min/μg).

*Statistics*

Data are presented as mean ± SEM. Data were analyzed by Student's *t*-test, one-way ANOVA followed by followed by followed by Bonferroni multiple comparison, as appropriate, using Prism 9 or later versions (GraphPad Software, San Diego, CA). Differences were considered statistically significant at P<0.05.

**Table S1: Major Proteomic hits from the hypothalamus before and after Ang-II treatment.** Numbers in bold highlight the effect of Ang-II treatment.

| Accession Number | Description [OS=Mus musculus] | Abundance Score | | | |
| --- | --- | --- | --- | --- | --- |
|  |  | Male | Male + Ang-II | Female | Female + Ang-II |
| O70481 | E3 ubiquitin-protein ligase UBR1 | 161.5 | **443.2** | 127.3 | **1811** |
| P46935 | E3 ubiquitin-protein ligase Nedd4 | 102.1 | 81.9 | 94.7 | 91.1 |
| Q8CFI0 | E3 ubiquitin-protein ligase Nedd4-like | 91.8 | 73.8 | 92.6 | 134.6 |
| Q8VE47 | Ubiquitin-like modifier-activating enzyme 5 | 84.6 | 124.8 | 84.7 | 104.7 |
| Q8K3W0-2 | BRISC and BRCA1-A complex member 2 | 157 | **101.7** | 111.4 | **90** |
| Q9CPX6 | ubiquitin-like-conjugating enzyme ATG3 | 85.3 | 114.6 | 99.8 | 105.2 |
| G5E870 | E3 ubiquitin-protein ligase TRIP12 | 87 | 116.3 | 80.4 | 100.5 |
| Q9CQY1 | Ubiquitin-like protein ATG12 | 60.1 | 78.9 | 79 | 74.7 |
| Q8CDG3 | Deubiquitinating protein VCIP135 | 103.8 | 134 | 94.8 | 93.9 |
| G5E870 | E3 ubiquitin-protein ligase TRIP12 | 116.3 | 87.0 | 100.5 | 80.4 |
| Q9D906 | Ubiquitin-like modifier-activating enzyme ATG7 | 103.1 | 110.6 | 118.2 | 105 |
| Q8C7R4 | Ubiquitin-like modifier-activating enzyme 6 | 100.9 | 103.5 | 97.8 | 106 |
| Q9D2M8 | Ubiquitin-conjugating enzyme E2 variant 2 | 104.8 | 106.9 | 110.3 | 97 |
| Q9Z1K5 | E3 ubiquitin-protein ligase ARIH1 | 78.2 | 79.8 | 80.4 | 81.8 |
| Q9JKB1 | Ubiquitin carboxyl-terminal hydrolase isozyme | 94 | 95.3 | 101.6 | 100.7 |
| Q9JI90-1 | E3 ubiquitin-protein ligase rnf14 | 84 | 101.2 | 98.7 | 88.3 |
| Q8C7M3-1 | E3 ubiquitin-protein ligase TRIM9 | 125.1 | 137.3 | 137.8 | 137.1 |
| Q6I6G8 | E3 ubiquitin-protein ligase HECW2 | 79.9 | 79 | 98.5 | 87.4 |
| P61089 | ubiquitin-conjugating enzyme E2 N] | 95.1 | 92.3 | 92.5 | 92.9 |

**Table S2: List of donated human cardiac samples.** Cardiac left ventricle samples were obtained from the Wisconsin Donor Network and Versiti Blood Center of Wisconsin organ procurement program (MCW) and Duke University Department of Surgery (DU).

| **Normotensive patients** | | | | | |
| --- | --- | --- | --- | --- | --- |
| **Patient ID** | **Source** | | **Race** | **Sex** | **Age** |
| 12364 | MCW | | Non-Hispanic Black | M | 35 |
| 14333 | MCW | | Non-Hispanic Black | M | 54 |
| 206 | DU | | Non-Hispanic Black | M | 29 |
| 94 | DU | | Non-Hispanic Black | F | 37 |
| 168 | DU | | Non-Hispanic Black | F | 28 |
| 13209 | MCW | | Caucasian | M | 45 |
| 79 | DU | | Caucasian | M | 41 |
| 150 | DU | | Caucasian | M | 52 |
| 77 | DU | | Caucasian | F | 44 |
| 85 | DU | | Caucasian | F | 49 |
| 13005 | MCW | | Caucasian | F | 36 |
| 12681 | MCW | | Caucasian | F | 51 |
| **Hypertensive patients** | |  |  |  |  |
| **Patient ID** | **Source** | | **Race** | **Sex** | **Age** |
| 13082 | MCW | | Non-Hispanic Black | M | 51 |
| 272 | DU | | Non-Hispanic Black | M | 55 |
| 144 | DU | | Non-Hispanic Black | M | 46 |
| 44 | DU | | Non-Hispanic Black | F | 49 |
| 97 | DU | | Non-Hispanic Black | F | 44 |
| 103 | DU | | Non-Hispanic Black | F | 42 |
| 12266 | MCW | | Non-Hispanic Black | F | 50 |
| 12917 | MCW | | Caucasian | M | 54 |
| 118 | DU | | Caucasian | M | 57 |
| 127 | DU | | Caucasian | M | 51 |
| 80 | DU | | Caucasian | F | 44 |
| 173 | DU | | Caucasian | F | 51 |

**Supplemental Figures and Legends**

**Figure S1**

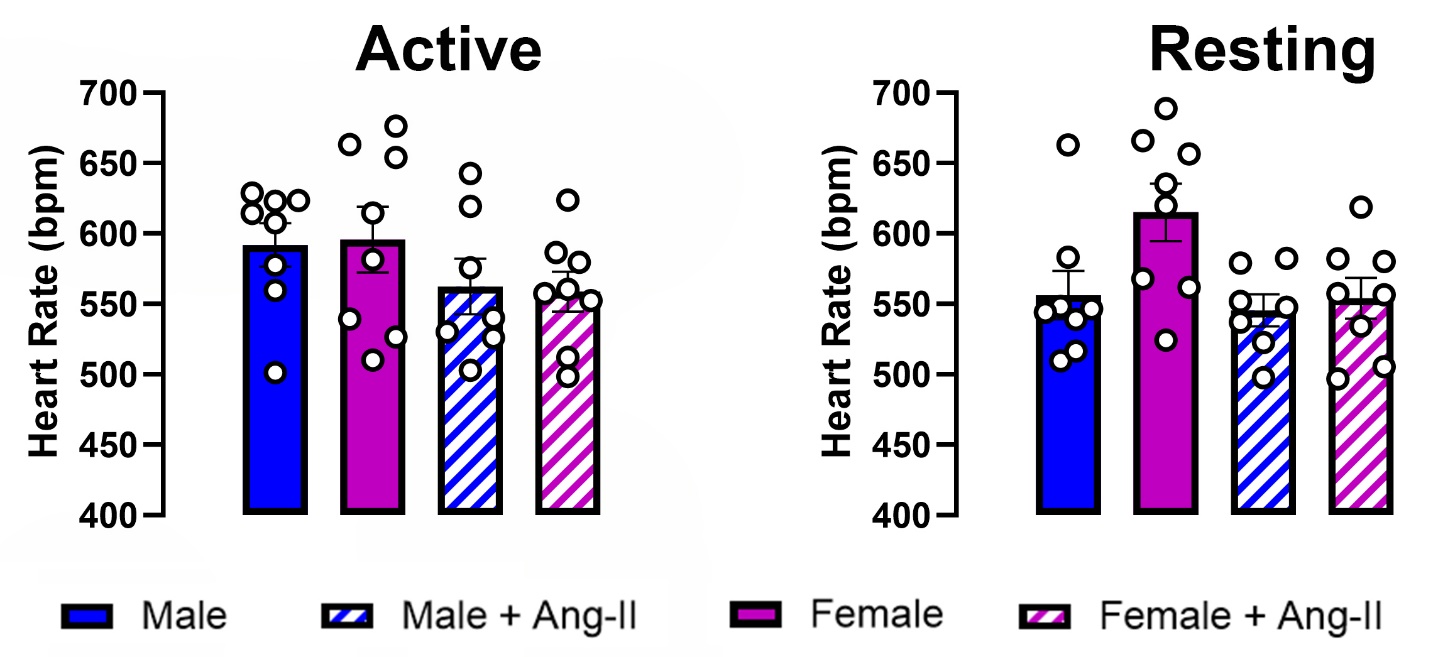

**Figure S1:** Neurogenic hypertension is sex-dependent in C57Bl6/J mice. Heart rate was recorded in C57Bl6/J mice at baseline (solid bars) and four weeks after Ang-II (600 ng/kg/min, hatched bars) infusion (n=7-8/group) during the active and resting. Data are shown as mean ±SEM.

**Figure S2**

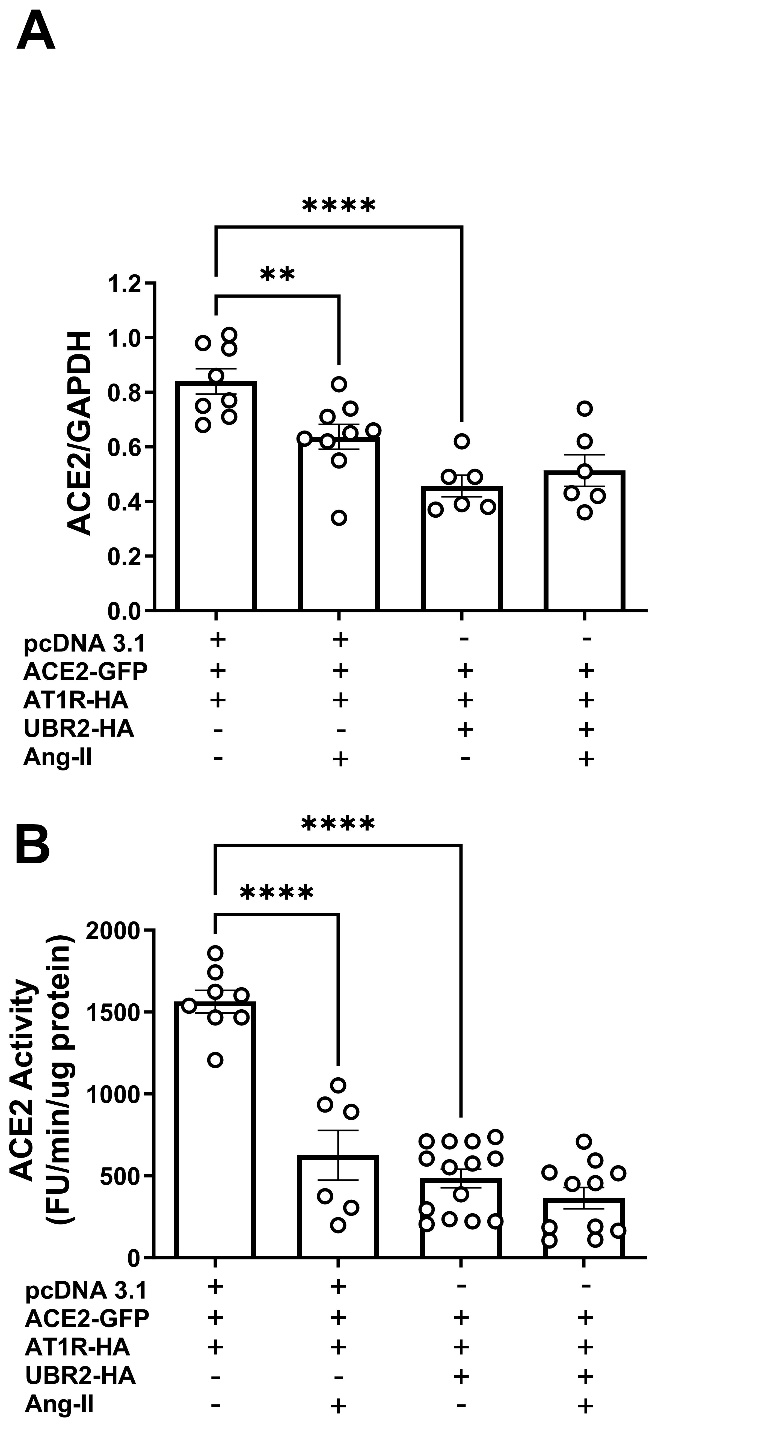

**Figure S2: Overexpression of UBR2 induces degradation of ACE2.** HEK293T cells were transfected with hACE2-GFP, AT_1_R-HA and UBR2-HA plasmids for 6 hours before exposure to Ang-II (100 nM) for 4 hours. Quantitative data for ACE2 protein expression based on densitometric analysis (**A**) and ACE2 activity (**B**) (n=6-9). Statistical significance: One-way ANOVA: **P<0.01 and ****P<0.0001.

**Figure S3**

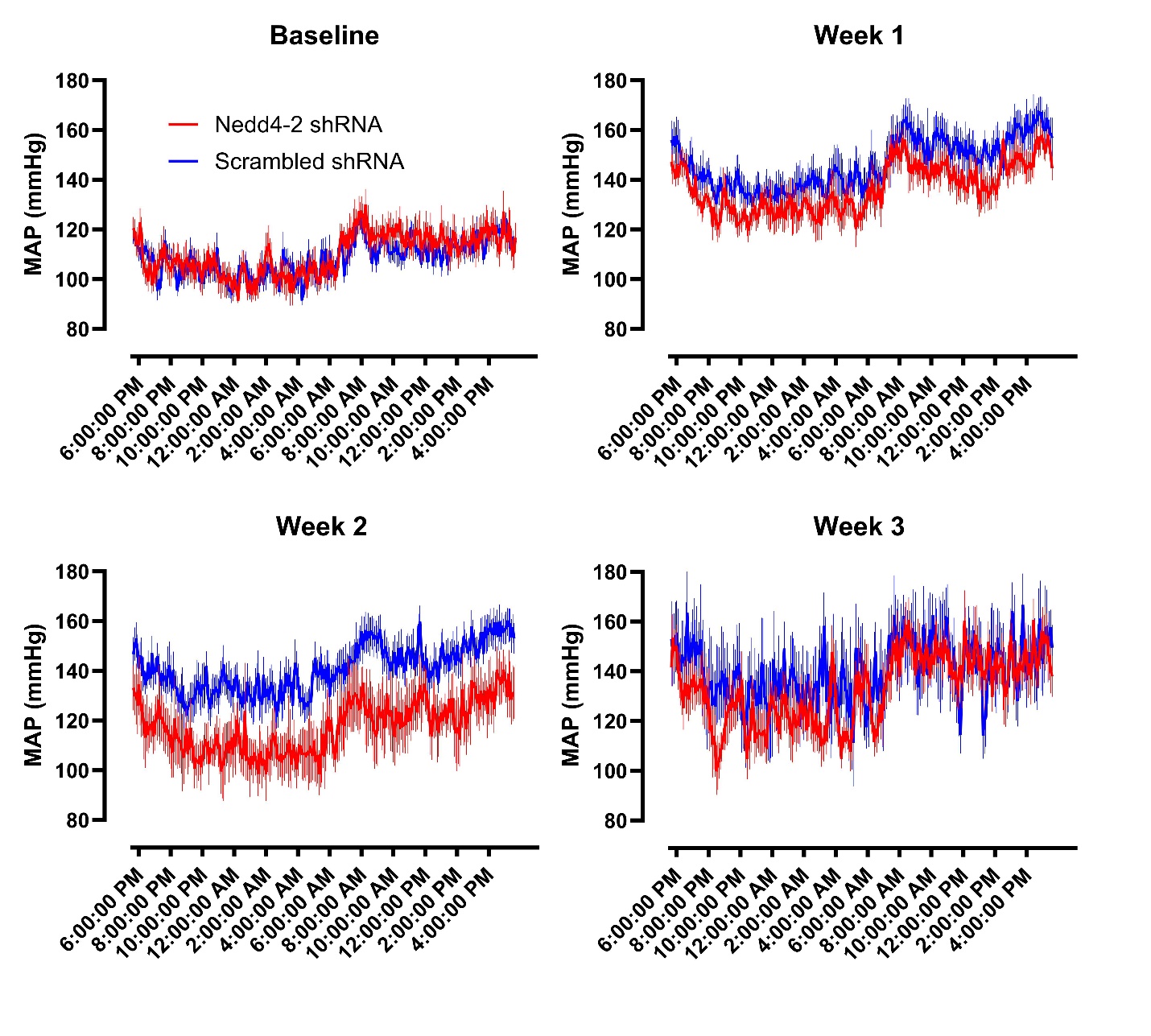

**Figure S3: Impact of Nedd4-2 knockdown in the brain.** Blood pressure recording for NEDD4-2 shRNA icv injected hypertensive male mice. MAP for UBR1 knockdown mice is represented against control mice treated with scrambled shRNA lentivirus in the PVN. Data is recorded for a 24 h period for 3 weeks following Ang-II infusion.
